# Suboptimal biological sampling as a probable cause of false-negative COVID-19 diagnostic test results

**DOI:** 10.1101/2020.05.05.20091728

**Authors:** Natalie N. Kinloch, Gordon Ritchie, Chanson J. Brumme, Winnie Dong, Weiyan Dong, Tanya Lawson, R. Brad Jones, Julio S.G. Montaner, Victor Leung, Marc G. Romney, Aleksandra Stefanovic, Nancy Matic, Christopher F. Lowe, Zabrina L. Brumme

## Abstract

Improper nasopharyngeal swab collection could contribute to false-negative COVID-19 results. In support of this, specimens from confirmed or suspected COVID-19 cases that tested negative or indeterminate (*i.e.* suspected false-negatives) contained less human DNA (a stable molecular marker of sampling quality) compared to a representative pool of specimens submitted for testing.

## Main text

Accurate COVID-19 diagnosis is critical to a successful clinical and public health response. Current COVID-19 tests detect one or more targets in the SARS-CoV-2 RNA genome, usually by real-time RT-PCR, and nasopharyngeal swabs have been the preferred sample for testing to date (1). While PCR-based tests are highly sensitive, the false-negative COVID-19 testing rate by nasopharyngeal swab (2) was nevertheless estimated to be >30% in a recent study (3), though the report in question had not yet undergone peer review. Factors other than the molecular technology contribute to test sensitivity, including the timing of sample collection with respect to infection stage (4) as well as specimen storage and transport (2). Improper specimen collection could also contribute to false negative COVID-19 test results. Although nasopharyngeal swabs are routinely ordered for respiratory viruses, the collection of a high quality specimen requires training and expertise as it involves insertion of the swab to posterior nasopharynx, a depth of roughly 7cm, followed by rotation and withdrawal of the swab (5). To investigate suboptimal sample collection as a possible cause of false negative test results, we quantified human DNA levels recovered on nasopharyngeal swabs submitted to a single laboratory for COVID-19 testing, hypothesizing that human DNA could serve as a stable molecular marker of specimen collection quality.

## Methods

The St. Paul’s Hospital (SPH) Virology laboratory in Vancouver, Canada is one of five provincially-designated SARS-CoV-2 diagnostic laboratories in British Columbia. COVID-19 testing on nasopharyngeal swabs (Copan UTM^®^ collection kit or BD universal viral transport system) was performed by total nucleic acid extraction from 500μL medium on the Roche MagnaPure 96 followed by real-time reverse-transcriptase (RT)-PCR on the Roche Lightcycler 480 using E-Sarbeco (6) and US-CDC RNAseP (7) primer/probe sets, or by testing on the Roche cobas^®^ 6800.

In March and April 2020 we identified 31 suspected false negative or indeterminate nasopharyngeal swab test results from presumed or confirmed COVID-19 cases for which >1mL medium remained for re-testing (henceforth referred to as “suspected false negative samples”). These included initial negative samples from individuals who subsequently tested positive within ≤12 days (N=11; range 0-12 days), samples from confirmed cases that returned indeterminate results (N=3; defined as detection of only 1 of 2 SARS-CoV-2 targets with a Cycle threshold [C_t_] value >35) and samples from individuals who repeatedly tested negative but for whom there was high clinical suspicion of infection by the responsible treating physician with no alternate diagnosis established (N=17). For comparison we tested a convenience sample of 87 consecutively submitted nasopharyngeal swabs for which >1mL medium remained. Remnant specimens were stored at −20°C until re-testing. To standardize nucleic acid extraction methodology across all specimens and to maximize SARS-CoV-2 RNA recovery and concentration, one milliliter of each remnant specimen was extracted on the BioMérieux EasyMag and eluted in 35μL buffer. SARS-CoV-2 detection in suspect false negative samples was re-attempted using a nested RT-PCR and sequencing protocol targeting conserved regions in ORF-1a and Spike (8). Human DNA levels were quantified using droplet digital PCR (ddPCR), a technique where each sample is fractionated into 20,000 nanolitre-sized water-in-oil droplets prior to PCR amplification with sequence-specific primers and fluorescent probes. Droplets are assayed at endpoint, after which Poisson statistics are used to calculate input template concentrations. The protocol was originally developed to quantify human genome copy numbers to high accuracy and to estimate DNA shearing in the sample (though the latter is not essential for the present purpose) (9). Briefly, the extract is combined with primer/probe sets targeting two regions in the human RPP30 gene ~9kb apart, ddPCR Supermix for Probes (no dUTPs, BioRad), XhoI restriction enzyme and nuclease free water. Primers and probes are: RPP30 Forward-GATTTGGACCTGCGAGCG, RPP30 Probe-VIC-CTGACCTGA-ZEN-AGGCTCT-3IABkFQ, RPP30 Reverse Primer-GCGGCTGTCTCCACAAGT; RPP30 Shear Forward Primer-CCAATTTGCTGCTCCTTGGG, RPP30 Shear Probe-FAM-AAGGAGCAA-ZEN-GGTTCTATTGTAG-3IABkFQ, RPP30 Shear Reverse Primer-CATGCAAAGGAGGAAGCCG (Integrated DNA Technologies; ZEN = internal ZEN™ quencher; 3IABkFQ = 3’ Iowa Black^®^ Fluorescent Quencher). Droplets were generated using the Automated Droplet Generator (BioRad) and cycled at 95°C for 10 minutes; 40 cycles of (94°C for 30 seconds, 53°C for 1 minute) and 98°C for 10 minutes. Droplets were analyzed on a QX200 Droplet Reader (BioRad) using QuantaSoft software (BioRad, version 1.7.4). Measured copies of RPP30/reaction are averaged across primer/probe sets, divided by two (as each human cell carries two RPP30 copies) and normalized to input volume to determine cells/μL of extract. A convenience panel of 94 remnant nucleic acid extracts performed on the Roche MagnaPure96 as part of COVID-19 testing were assessed for human RNAseP RNA levels using the US-CDC protocol on a Roche Lightcycler 480 (7). This study was approved by the Providence Health Care/University of British Columbia and the Simon Fraser University Research Ethics Boards.

## Results

We began by re-testing the 31 suspected false negative specimens using a nested RT-PCR and confirmatory sequencing protocol targeting conserved regions in ORF-1a and Spike (8), reasoning that nested PCR could offer increased SARS-CoV-2 detection sensitivity. Before implementing this protocol we performed a blinded validation on a test panel of 24 EasyMag extracts of SARS-CoV-2 negative and positive samples (where positives yielded C_t_ values of 21.5 to 35.4 on the LightCycler 480) and confirmed 100% concordance with nested RT-PCR/sequencing results (not shown). All 31 suspect false negatives again tested negative by nested RT-PCR except one that yielded an amplicon for ORF1a but not Spike (this was one of the three indeterminate results by real-time RT-PCR). Based on this result, this sample was subsequently excluded from downstream investigation. This indicated that the original negative results were not likely attributable to suboptimal performance of the real-time RT-PCR assay, but rather suggested that SARS-CoV-2 RNA was exceedingly low or absent in these samples.

We then investigated whether human DNA recovered on the nasopharyngeal swab could serve as a molecular marker of specimen collection quality, reasoning that DNA (by virtue of its stability) would be well-preserved in remnant clinical specimens. We employed a sensitive, multiplexed ddPCR protocol for absolute human RPP30 gene copy number quantification (9). Overall, we observed significantly lower human DNA levels in suspected false negative nasopharyngeal swab samples compared to a panel of consecutive samples submitted for testing during the same period, though overlap between groups was still substantial (Figure 1, p<0.001). Specifically, suspected false negative specimens harbored a median 3,409 (Interquartile range [IQR]: 1,229- 4,295) human cells/μL of extract while comparison samples harbored a median 5,539 (IQR 3,649- 7,744) cells/μL. This strongly supports suboptimal biological sampling as a probable cause of false-negative COVID-19 test results.

**Figure 1:**
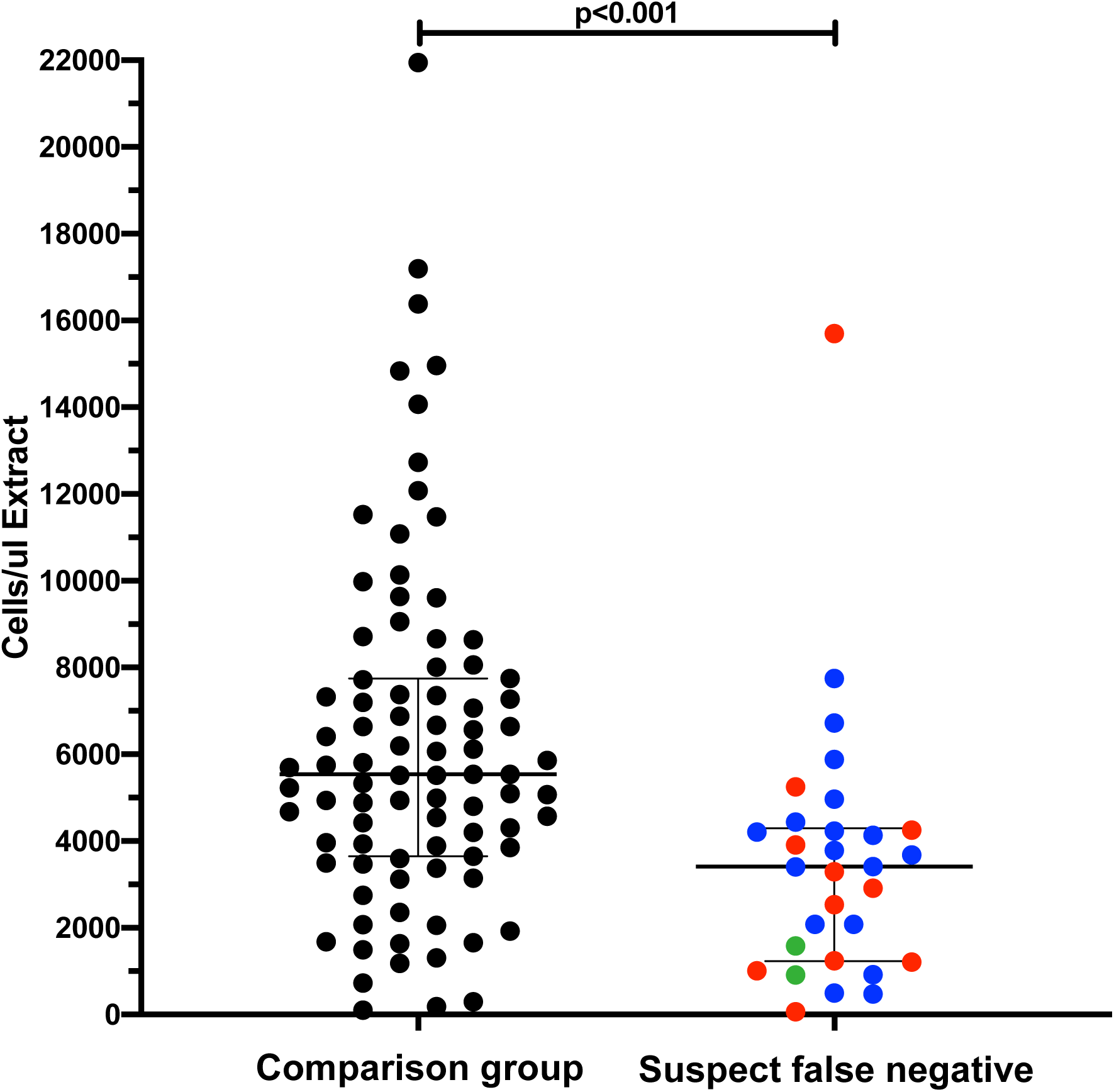
Suspected false negative COVID-19 test samples harbored significantly lower human DNA levels compared to a representative pool of specimens submitted for testing. Human DNA levels (RPP30 gene target) were measured using ddPCR in nasopharyngeal extracts as a molecular marker of biological sampling quality. “Suspect false negatives” included samples from individuals who subsequently tested positive < 14 days later (**red**), samples from known cases that yielded indeterminate results (**green)** and samples from individuals with high clinical suspicion of being infected but never clinically confirmed (**blue**). Note that there are only two samples in the indeterminate category, as the third re-tested partially positive by nested RT-PCR, and therefore was excluded from human DNA analysis. The comparison dataset was a consecutive set of 87 samples submitted for testing in April 2020 to the same laboratory.

## Discussion

Our results underscore the importance of proper training and technique in the collection of high quality nasopharyngeal specimens. They also highlight the potential utility of including a molecular marker of sampling quality in SARS-CoV-2 diagnostic RT-PCR assays. While the major commercial diagnostic assays include internal controls, these do not necessarily measure specimen quality. In the Roche cobas^®^ SARS-CoV-2 assay for example (https://www.fda.gov/media/136049), an armored RNA construct is spiked into specimens. Its subsequent detection confirms the validity of nucleic acid extraction and RT-PCR amplification, but does not provide a measure of biological sampling quality. The US CDC COVID-19 test includes a human RNAseP RNA-specific primer/probe set to assess sampling quality, but the results are interpreted qualitatively (7). Specifically, the most recent US-CDC’s instructions for use (issued 15-Mar-2020) states that failure to detect RNAseP within 40 PCR cycles can denote improper assay set up, poor nucleic RNA yield/quality after extraction, and/or the absence of sufficient human cellular material in the sample due to poor specimen collection or storage. As such, specimens for which *neither* SARS-CoV-2 nor RNAseP are detected are considered invalid. To estimate the range of RNAseP RNA content in clinical samples we retrospectively tested a panel of 94 SARS-CoV-2 nucleic acid extracts generated on the MagnaPure 96. Despite storage at 4°C since extraction, their RNAseP RNA C_t_ values still ranged from 19.65 to 31.68, where the 90th percentile C_t_ value was 26.10. Thus, even the lowest decile of samples in terms of RNAseP RNA levels (possibly representing those for which sampling was the least robust) still amplified well before C_t_<40, suggesting that this threshold may be insufficient to assess biological sampling quality. It is important to note however that our study was not designed to identify a threshold of human DNA (or RNA) that could define a properly-collected SARS-CoV-2 nasopharyngeal swab. Future studies attempting to do so would need to consider that recovery efficiency (and thus total yield) of different types of nucleic acid may differ by extraction platform, and possibly by swab type, so thresholds may need to be assay specific.

## Conclusion

Our observations strongly support suboptimal biological sampling, but not PCR sensitivity for SARS-CoV-2 RNA detection, as a contributing cause of false-negative COVID-19 test results.

## Data Availability

Raw data are available upon request to corresponding authors.

## Acknowledgement

We thank the laboratory teams at the St. Paul’s Hospital Virology Laboratory and the BC Centre for Excellence in HIV/AIDS for technical assistance. We thank Dr. Christopher Sherlock for helpful discussions.

